# Target enrichment improves culture-independent detection of *Neisseria gonorrhoeae* direct from sample with Nanopore sequencing

**DOI:** 10.1101/2024.01.09.24301039

**Authors:** Teresa L. Street, Nicholas D. Sanderson, Leanne Barker, James Kavanagh, Kevin Cole, The GonFast Investigators Group, Martin Llewelyn, David W. Eyre

**Author notes:** **Corresponding author and email address:** Teresa L. Street. These authors contributed equally to this manuscript (Teresa L. Street, Nicholas D. Sanderson). Author order was determined by mutual agreement. **Repositories** European Nucleotide Archive study code PRJEB64347.

## Abstract

Multi-drug resistant *Neisseria gonorrhoeae* infection is a significant public health risk. Rapidly detecting *N. gonorrhoeae* and antimicrobial resistant (AMR) determinants by metagenomic sequencing of urine is possible, although high levels of host DNA and overgrowth of contaminating species hamper sequencing and limit *N. gonorrhoeae* genome coverage. We performed Nanopore sequencing of nucleic acid amplification test-positive urine samples and culture-positive urethral swabs with and without probe-based target enrichment, using a custom SureSelect panel to investigate selectively enriching for *N. gonorrhoeae* DNA. Probes were designed to cover the entire *N. gonorrhoeae* genome, with 10-fold enrichment of probes covering selected AMR determinants. Multiplexing was tested in subset of samples. The proportion of sequence bases classified as *N. gonorrhoeae* increased in all samples after enrichment, from a median (IQR) of 0.05% (0.01-0.1%) to 76% (42-82%), giving a corresponding median improvement in fold genome coverage of 365-times (112–720). Over 20-fold coverage, required for robust AMR determinant detection, was achieved in 13/15 (87%) samples, compared to 2/15 (13%) without enrichment. The four samples multiplexed together also achieved >20x genome coverage. Coverage of AMR determinants was sufficient to predict resistance where present, and genome coverage also enabled phylogenetic relationships to be reconstructed. Probe-based target enrichment can improve *N. gonorrhoeae* genome coverage when sequencing DNA extracts directly from urine or urethral swabs, allowing for robust detection of AMR determinants. Additionally, multiplexing prior to enrichment provided enough genome coverage for AMR detection and reduces the costs associated with this method.

**Impact statement:** *Neisseria gonorrhoeae* infection presents a significant public health risk, with multi-drug resistance present globally. Early detection helps control the spread of antimicrobial resistant strains. Genome sequencing can be used to detect infections in samples collected directly from patients, without the need to grow any microorganisms in a laboratory first, and this has already been demonstrated for gonorrhoea using urine samples. With enough sequence information it is also possible to detect antimicrobial resistance (AMR), allowing both detection of infection and information on treatment choices from the same test. This study assesses a method for enriching *N. gonorrhoeae* sequence data from urine and urethral swabs, and analyses the impact of enrichment on the detection of genes known to cause antibiotic resistance. We show that enriching for *N. gonorrhoeae* DNA prior to sequencing can improve the detection of some AMR genes, and by testing several samples at the same time we can reduce the costs associated with this method.

**Data summary:** The sequence data generated in this study are deposited in the European Nucleotide Archive (ENA, https://www.ebi.ac.uk/ena/browser/) and are publicly available under study code PRJEB64347. The authors confirm all supporting data, code and protocols are provided within the article or through supplementary data files.

## 6. Introduction

Multi-drug resistant *Neisseria gonorrhoeae* infection presents a significant global public health risk(1), with resistance to ceftriaxone and/or azithromycin detected in countries worldwide(2–4). Additionally, cases of *N. gonorrhoeae* infection are increasing. In 2019, 117,881 cases were reported in the EU/EEA, over half of which were reported in the UK with 116 cases/100,000 population(5). Early detection and effective treatment is vital to prevent the spread of drug-resistant strains.

Metagenomic sequencing (mNGS) directly from clinical samples offers the potential to reduce the time to diagnosis and can provide additional information such as detection of antimicrobial resistance(6), particularly useful in settings where culture is not routine. mNGS has previously been used to both identify *N. gonorrhoeae*(7) and detect antimicrobial resistance (AMR) determinants(8) directly from urine samples. A limitation of mNGS is the high levels of contaminating human DNA present in extracts from clinical samples which limits the amount of pathogen sequence data generated(6). Target enrichment is a technique that allows enrichment of genomes or genomic regions of interest prior to sequencing: Biotinylated RNA oligonucleotide probes, designed to target specific sequences, allow capture and enrichment of these regions of interest by hybridisation and magnetic pulldown. Target enrichment has previously been demonstrated to improve the amount of pathogen sequence generated from mNGS(9–11), improving genome coverage and detection of AMR determinants(12).

This study assesses the ability of probe-based target enrichment to enrich for *N. gonorrhoeae* DNA directly from urine and urethral swab samples for the detection of AMR determinants using the Oxford Nanopore Technologies (ONT) sequencing platform. We also tested the efficiency of target enrichment after multiplexing samples together, aiming to reduce the costs associated with both probe-based enrichment and sequencing.

## 7. Methods

### 7.1 Samples

Samples were selected from those collected as part of a wider study, conducted with NHS Research Ethics approval (reference 19/EM/0029). Participants were recruited at sexual health clinics at Oxford University Hospitals NHS Trust, UK, and University Hospitals Sussex NHS Trust, Brighton, UK. Male patients presenting with symptomatic urethritis were eligible to participate and were recruited following informed consent. In Brighton, urine samples were collected into universal tubes containing boric acid (Medline Scientific) for stabilization during transportation to Oxford. In Oxford, samples were collected into universal tubes without boric acid. Urethral swabs were placed into Sigma VCM preservation medium (MWE). All samples were tested for *N. gonorrhoeae* using the BD Viper system (Becton Dickinson), with confirmatory testing for *N. gonorrhoeae* undertaken using the BD Max platform (Oxford) or with the FTD gonorrhea confirmation NAAT assay (Fast Track Diagnostics) (Brighton). Antimicrobial susceptibility testing of cultured bacteria from urethral swabs was performed manually according to EUCAST methods by disc diffusion (for nalidixic acid, penicillin, azithromycin, cefuroxime and ciprofloxacin) and E-tests (for cefixime and cefotaxime) in Brighton, and by E-tests (for ciprofloxacin, azithromycin and ceftriaxone) in Oxford.

### 7.2 Custom probe capture library design

A SureSelectXT custom probe library was designed with Agilent, the manufacturer, to cover 17 closed genomes of *N. gonorrhoeae* identified by the authors to represent the diversity present across the *N. gonorrhoeae* species (Table S1). Additionally, probes designed to cover genes known to confer antimicrobial resistance in *N. gonorrhoeae* were represented 10-fold in the final library (Table S2). The final probe library consisted of consisting of approximately 50,000 120mer RNA probes.

### 7.3 DNA extraction, library preparation and sequencing

DNA was extracted from nucleic acid amplification test (NAAT)-positive urine samples and urethral swabs using the QIAamp UCP pathogen mini kit (Qiagen), as previously described(7). Samples without enrichment were prepared for sequencing on version 9.4.1 flow cells following library preparation with the rapid PCR barcoding kit (SQK-RPB004, ONT) also as previously described(7). One sample was run per flow cell on a GridION sequencer. For SureSelect enrichment, samples were prepared following the Sequence Capture protocol (ONT), using the ligation kit (SQK-LSK110, ONT) for sequencing with a single sample per flow cell.

### 7.4 Preparation for multiplexed sequencing

To test the feasibility of multiplexing, 4 samples were barcoded with a modified protocol using the rapid PCR barcoding kit (SQK-RPB004, ONT). Briefly, 200ul DNA extract was fragmented by tagmentation in a 10 µl reaction with 2.5 µl FRM, then barcoded by PCR in a double volume reaction for 17 cycles with a 5-minute elongation time at 65°C. Post-PCR, barcoded DNA was processed following a modified Sequence Capture protocol, as follows. DNA was end-repaired and PCR adapters ligated as per the original protocol. The initial PCR was omitted, and instead equimolar amounts of each barcoded, amplified sample were pooled together prior to hybridisation with the custom probes. Post-hybridisation, the Sequence Capture protocol was followed as described by ONT and enriched DNA was prepared for sequencing using the ligation kit, as above.

### 7.5 Illumina sequencing of cultured isolates

Cultures from those samples with a viable stock were prepared for sequencing on an Illumina MiniSeq sequencer. *N. gonorrhoeae* DNA was extracted from cultured cells grown overnight on chocolate agar at 37°C with 5% CO_2_ using the QuickGene DNA tissue kit (Kurabo) on the QuickGene-Mini80 system (MP Biomedicals), and prepared for sequencing with the Nextera XT DNA library prep kit (Illumina).

### 7.6 Bioinformatic analysis

Nanopore sequences were base called and demultiplexed, where necessary, using Guppy (ONT, Version 5.0 or higher) automatically on the GridION sequencer. Sequencing reads classified as human were removed at source with the CRuMPIT workflow, as previously described(13). Sequences were mapped to the NC_011035.1 *N. gonorrhoeae* reference genome using minimap2 (v2.24-r1122) and variant called with clair2 (v2.1.1). Variants were filtered with a random forest classifier, and antimicrobial resistance genes were characterised from consensus genome sequences or assembled plasmid sequences, both as previously described(8). The Nextflow workflow is available at https://gitlab.com/ModernisingMedicalMicrobiology/genericbugontworkflow.

Illumina sequences from cultured isolates were mapped to the NC_011035.1 reference genome with SNIPPY(14) v4.6.0 to generate SNP-only consensus sequences. Positions with depth lower than 10x coverage were masked as Ns. Hence, both Nanopore and Illumina data were mapped to the same reference genome and consensus sequences generated from ONT or Illumina-specific variant calling pipelines. Both Illumina and ONT consensus genomes were masked for repeat regions using a self-self blast, with a minium window of 200bp and 90% identity score.

A phylogenetic tree was constructed using IQ-TREE v2.1.4-beta(15) and adjusted for recombination events using clonalframeML v1.12(16) using the runlistcompare wrapper script v0.3.8 (https://github.com/davideyre/runListCompare) for samples with suffficient genome coverage.

## 8. Results

### 8.1 Samples summary

Twelve urine samples and four urethral swabs were selected for use in this study. Eleven (11/12, 92%) urines were NAAT-positive for *N. gonorrhoeae* and one (1/12, 8%; sample 382UB) was NAAT-negative with a corresponding culture-positive urethral swab. All four (100%) urethral swabs were culture-positive for *N. gonorrhoeae*, with corresponding urines also NAAT-positive. A sample summary is provided in Figure 1 and Figure S1.

**Figure 1.**
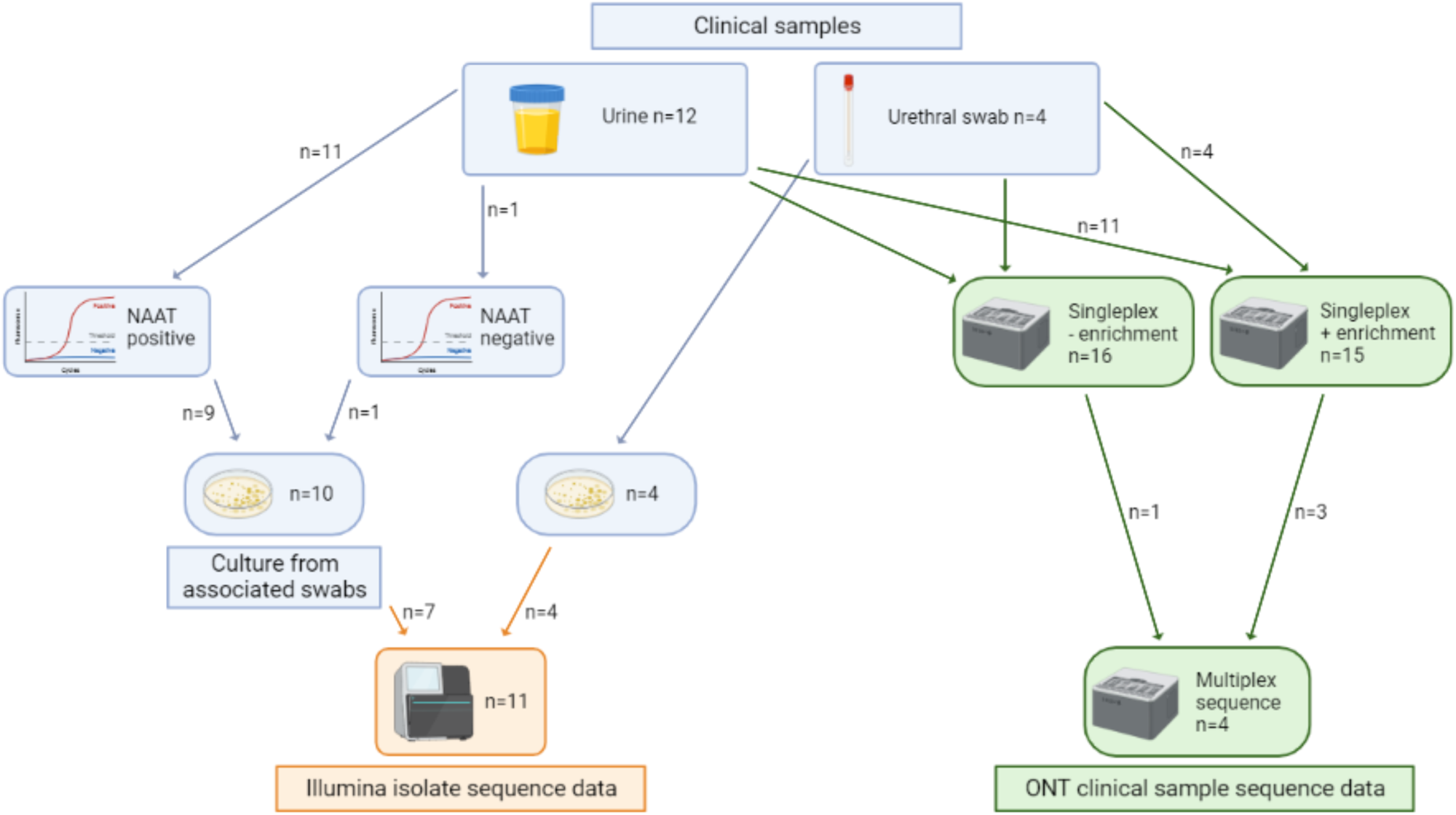
Sample summary. Figure created with BioRender

A total of 9/11 (82%) NAAT-positive urines had an additional corresponding culture-positive urethral swab, with 2/11 (18%) being culture-negative. Antimicrobial susceptibility testing results on cultured isolates were used for comparisons with mNGS-based predictions. Results were available for 14 urethral swabs (13 tested in Brighton and 1 in Oxford; 9 with NAAT-positive urines that were sequenced, 1 with a NAAT-negative urine that was sequenced, and 4 where the urethral swab was sequenced). All were sensitive to both cephalosporins and azithromycin (Table 1). For quinolone susceptibility, the single sample from Oxford (80U) was sensitive to ciprofloxacin, and of the Brighton samples 3/13 (23%) were sensitive and 10/13 (77%) resistant to nalidixic acid (Table 1). Brighton additionally tested for susceptibility to penicillin (sensitive in 2/13 (15%)) and for the presence of beta-lactamase (detected in 9/13, 69%).

**Table 1.** Antimicrobial susceptibility results. S, sensitive; I, intermediate; R, resistant; *for B-lactamase, N denotes susceptible; P denotes possible resistance.

| Sample | Sample type for culture | Sample type for sequencing | Ciprofloxacin/<br>Nalidixic acid | Cephalosporins | Azithromycin | Penicillin | B-lactamase* |
| --- | --- | --- | --- | --- | --- | --- | --- |
| 80U | Urethral swab | Urine | S | S | S | no test | no test |
| 202D | Urethral swab | Urethral swab | S | S | S | S | N |
| 265UB | Urethral swab | Urine | R | S | S | R | P |
| 301D | Urethral swab | Urethral swab | R | S | S | I | N |
| 303D | Urethral swab | Urethral swab | R | S | S | R | P |
| 304D | Urethral swab | Urethral swab | S | S | S | I | N |
| 305UB | Urethral swab | Urine | R | S | S | R | P |
| 318UB | No growth on culture | Urine |  |  |  |  |  |
| 321UB | Urethral swab | Urine | S | S | S | S | N |
| 342UB | Urethral swab | Urine | R | S | S | R | P |
| 347UB | Urethral swab | Urine | R | S | S | R | P |
| 358UB | Urethral swab | Urine | R | S | S | R | P |
| 361UB | No growth on culture | Urine |  |  |  |  |  |
| 364UB | Urethral swab | Urine | R | S | S | R | P |
| 367UB | Urethral swab | Urine | R | S | S | R | P |
| 382UB | Urethral swab | Urine | R | S | S | R | P |

### 8.2 Effect of enrichment on *Neisseria gonorrhoeae* genome coverage

Probe enrichment increased the proportion of *N. gonorrhoeae* sequencing reads compared to no enrichment and did not affect the total sequencing output of the runs (Figure 2). A comparison of total bases, bases classified as *N. gonorrhoeae* plus genome coverage breadth and depth before and after enrichment is provided in Table 2. The total number of bases per sample was similar between enriched (median 10.9Gb, IQR 8.1-17.3) and without enrichment (median 9.4Gb, IQR 8.4-10.8). Multiplexing samples onto a single flow cell reduced the number of bases per sample, as expected (median 478Mb, IQR 321-918) (Figure 2a). The median proportion of bases classified as *N. gonorrhoeae* increased from less than 0.5% of total sample bases without enrichment (median 0.05%, IQR 0.01-0.1) to more than 75% with enrichment (median 76%, IQR 42-82). Probe enrichment with multiplexing generated a lower median proportion of *N. gonorrhoeae* bases (33%, IQR 26-45) (Figure 2b). Average coverage depth of aligned reads increased from a median 5-fold (IQR 4.4-6.7) without enrichment to 2405-fold after enrichment (IQR 492-5423) and 63-fold (IQR 36-153) after multiplexed enrichment (Figure 2c). This resulted in all but two samples enriched with SureSelect probes achieving >96.5% of their genome sequenced at a depth of 300-fold or more (Table 2). Comparison of genome coverage with and without enrichment demonstrates no evidence that any bias was introduced by the Sequence Capture protocol (Figure 3).

**Figure 2.**
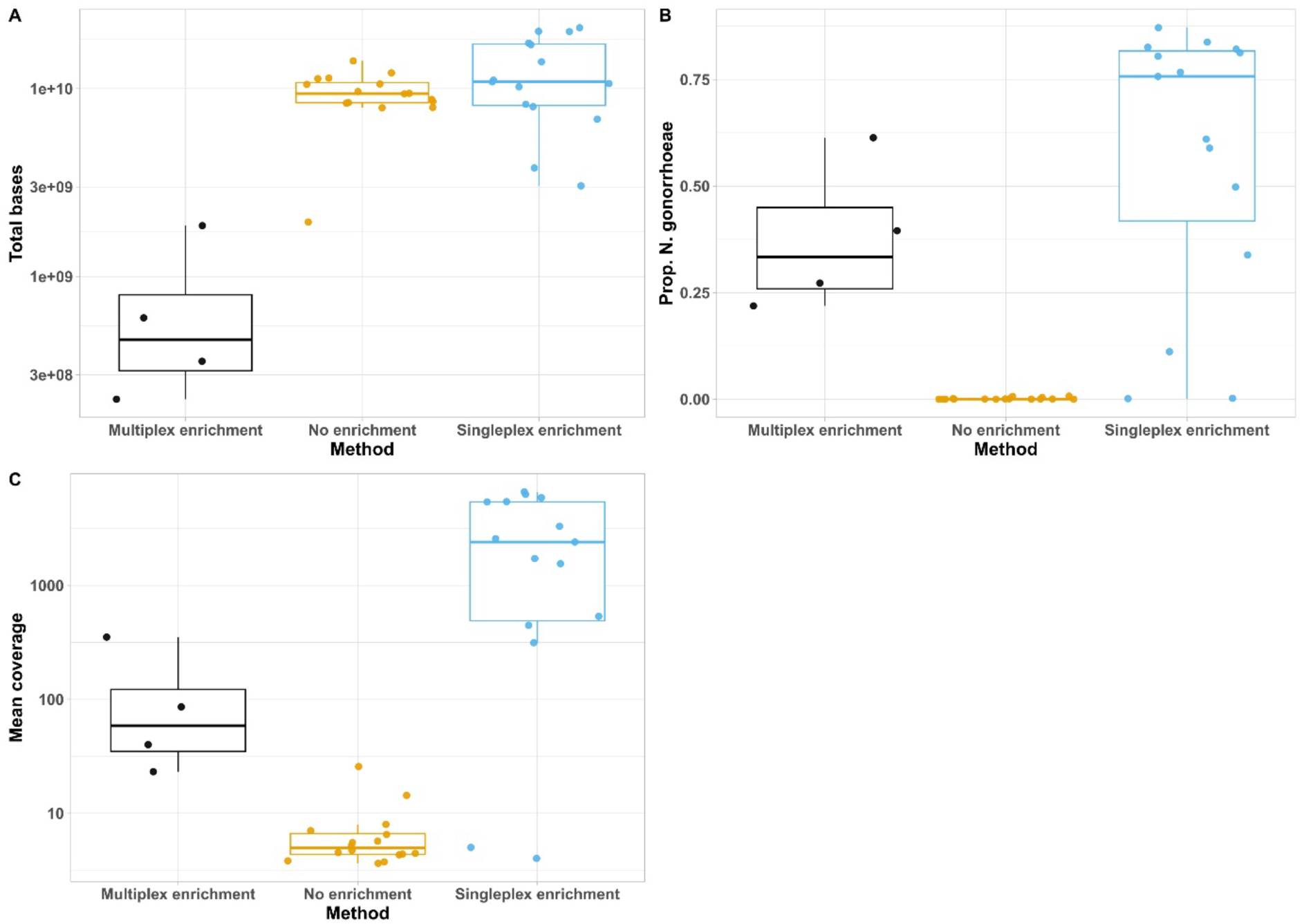
Comparison of sequence data before and after enrichment. Total number of bases per sequencing run (A); Percentage of bases classified as *N. gonorrhoeae* (B); Average coverage depth of *N. gonorrhoeae* genome per sequencing run (C) for samples sequenced without enrichment (orange), with singleplex enrichment (blue) or with enrichment in a multiplex of 4 samples (black).

**Figure 3.**
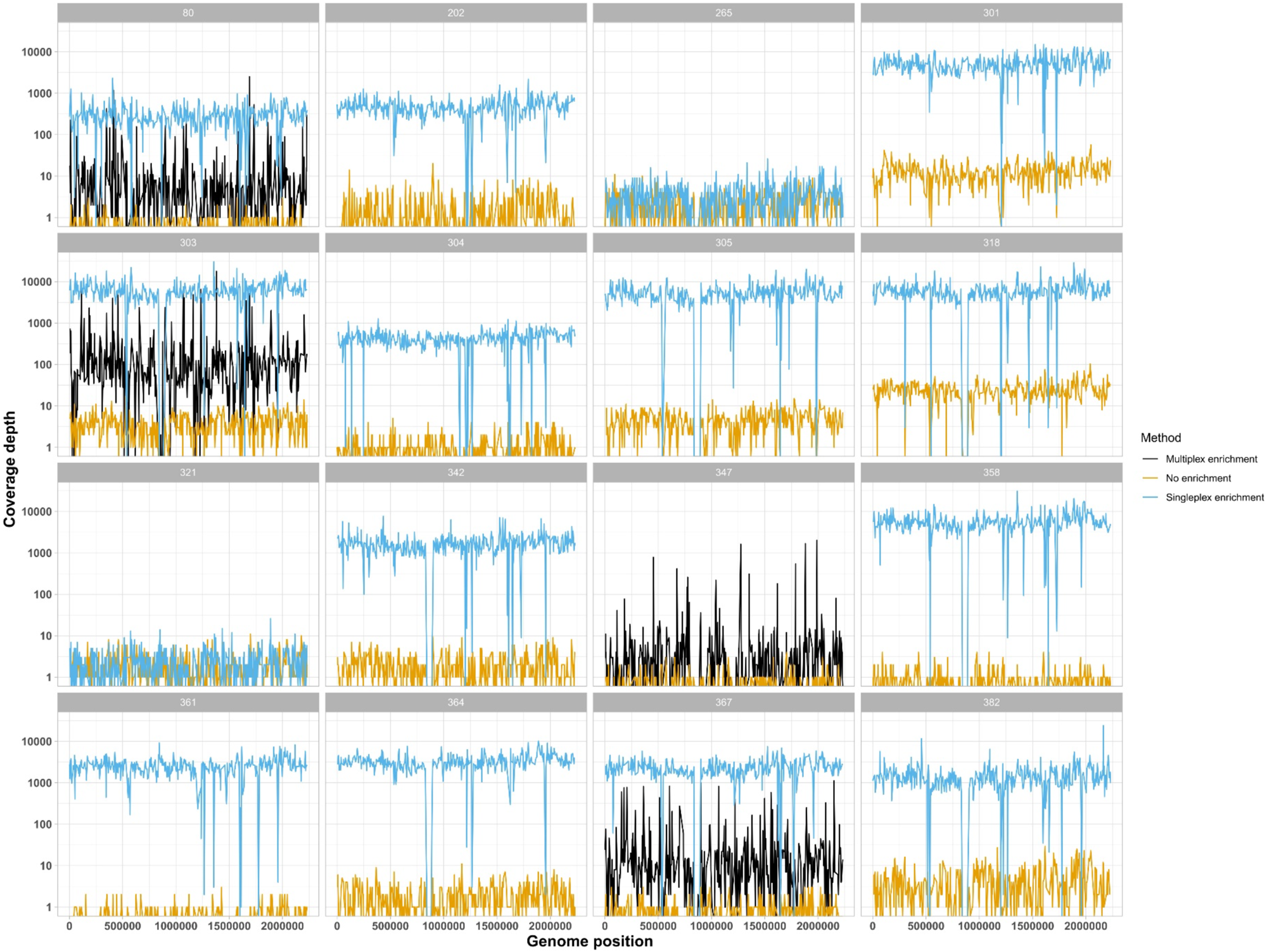
Genome coverage depth for samples sequenced without enrichment (orange), with singleplex enrichment (blue) or with enrichment in a multiplex of 4 samples (black). Sample 347 has sequence data without enrichment and with multiplexed enrichment only. Short regions of low coverage seen in blue represent areas of divergence between the sample and the reference genome used for mapping and/or the genomes included in the probe set. The similar troughs seen with the orange and blue lines suggest that divergence from the reference genome is the most likely explanation, rather than a gap in the probe set.

**Table 2.** Summary of bases classified as *N. gonorrhoeae* (Ngon), total bases, single-fold genome coverage breadth, and average genome coverage depth for all samples before and after singleplex enrichment.

|  | Without enrichment |  |  |  | With singleplex enrichment |  |  |  |  |
| --- | --- | --- | --- | --- | --- | --- | --- | --- | --- |
| Sample name | Ngon bases | Total bases | Genome coverage breadth (%) | Average coverage depth | Ngon bases | Total bases | Genome coverage breadth (%) | Average coverage depth | Coverage depth fold-enrichment, singleplex vs no enrichment |
| 80U | 749179 | 8434029003 | 3.46 | 7.98 | 1280208433 | 3782522608 | 97.44 | 314.12 | 39.3 |
| 202D | 4184726 | 9656053514 | 35.73 | 4.45 | 1550689332 | 13895666698 | 98.53 | 536.27 | 120.6 |
| 265UB | 5949401 | 11386449183 | 58.55 | 3.82 | 17922316 | 8014502433 | 73.45 | 5.01 | 1.31 |
| 301D | 35361341 | 7933536503 | 98.83 | 14.36 | 14947146476 | 17150934403 | 99.80 | 5413.55 | 377.0 |
| 303D | 9692745 | 9439958602 | 82.96 | 4.73 | 17664552591 | 21087141427 | 97.08 | 6637.14 | 1403.8 |
| 304D | 1937846 | 7906566477 | 17.88 | 4.32 | 1790026178 | 3038176523 | 97.49 | 447.24 | 103.5 |
| 305UB | 12659589 | 9391830588 | 86.68 | 5.53 | 14039629756 | 17449531128 | 96.75 | 5432.74 | 983.0 |
| 318UB | 62722932 | 8544860509 | 96.36 | 25.72 | 16557094125 | 20163378663 | 97.03 | 6295.03 | 244.8 |
| 321UB | 7745538 | 12127667362 | 57.53 | 4.38 | 9600291 | 6880830294 | 53.10 | 4.02 | 0.92 |
| 342UB | 4929172 | 14056225894 | 51.44 | 3.75 | 6489125469 | 10636014129 | 96.70 | 1724.94 | 460.4 |
| 358UB | 1399857 | 11292620134 | 10.46 | 5.22 | 15428552493 | 20118811661 | 97.09 | 5905.35 | 1131.6 |
| 361UB | 698401 | 8371283598 | 3.78 | 7.05 | 7748292558 | 10234243279 | 99.04 | 2576.51 | 365.4 |
| 364UB | 4923268 | 8765729938 | 51.80 | 3.64 | 8974767733 | 10869934865 | 96.84 | 3304.73 | 908.2 |
| 367UB | 1474533 | 10579314587 | 12.82 | 4.52 | 6713863610 | 8261193715 | 96.63 | 2405.44 | 532.2 |
| 382UB | 12826754 | 1949351698 | 71.43 | 6.52 | 5518490118 | 11085690959 | 96.73 | 1552.06 | 238.2 |

Enrichment failed for two samples (265UB, 321UB). Genome coverage breadth and depth were similar before and after enrichment: 59% breadth and 3.8-fold depth before and 73% breadth and 5-fold depth after for sample 265UB, and 58% breadth and 4.4-fold depth before and 53% breadth and 4-fold depth after for sample 321UB. Due to the high cost of reagents and the need to order these in batches, we were unable to repeat these experiments. A detailed summary of all sequence data generated is provided in Table S3.

### 8.3 Detection of antimicrobial resistance determinants

Our previous study determined that a minimum 20-fold genome coverage depth is required for robust AMR prediction(8). In this study 13/15 (87%) of samples achieved this after enrichment, with a minimum coverage depth of 314-fold (Table S4). Sequence data was interrogated for variants within genes known to confer resistance to macrolides, quinolones, cephalosporins, and for the presence of plasmid-mediated genes conferring resistance to penicillin and tetracycline (Table S4). None of the samples in this study were phenotypically resistant to cephalosporins or azithromycin, and no genetic determinants known to confer resistance to these were detected, despite >140-fold coverage of *mtrR* and >90-fold coverage of *penA*, *ponA* and *porB* in those samples with successful enrichment (Figure 4). Ten (10/16, 63%) samples were reported as resistant to nalidixic acid. Mutations in *gyrA* (S91F, D95G) were detected in two samples before enrichment. After enrichment, *gyrA* was sequenced to a depth of at least 115-fold and mutations were detected in 9/9 (100%) of those samples with singleplex enrichment: singleplex enrichment sequence data was not available for one sample (347UB). Additionally, *gyrA* mutations S91F and D95G were found in sample 361UB after enrichment, which did not grow in culture and therefore had no susceptibility data reported by the microbiology laboratory.

**Figure 4.**
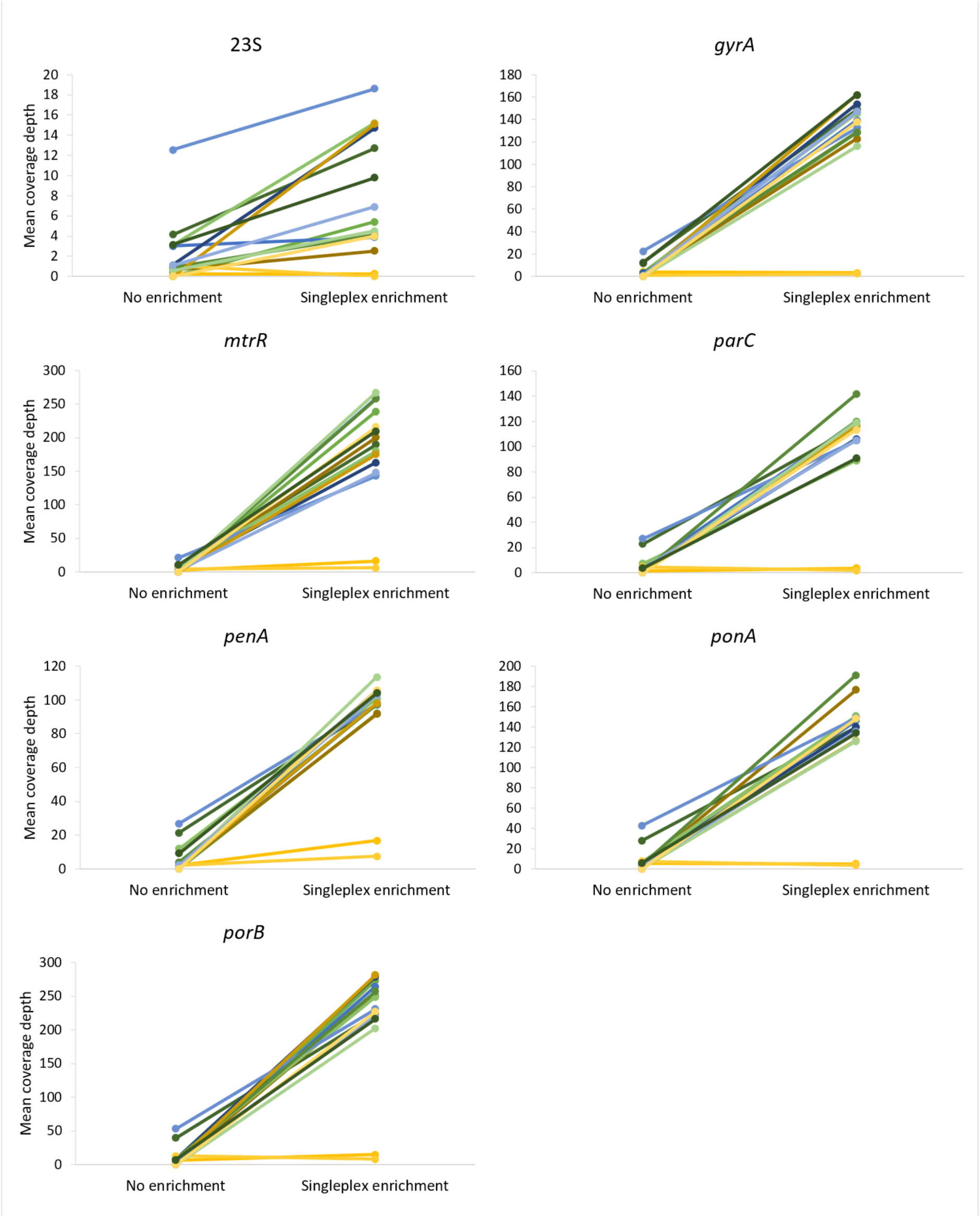
Comparison of mean coverage depth for 7 antimicrobial resistance determinants before and after target enrichment. Different colours are used for each sample pair with and without enrichment.

For plasmid-mediated resistance, no sequence data containing *ermB*, *ermC*, *ermF* or *mef* was detected, corresponding with laboratory results of susceptibility to azithromycin. Beta-lactamase was detected in 9/16 (56%) samples by the laboratory, and sequencing without enrichment detected the presence of *blaTEM-1* in 6 of these samples, and additionally in sample 361UB which did not have susceptibility data reported After enrichment, *blaTEM-1* was only detected in 2 samples using the criteria defined previously(8), and in 3 samples when considering coverage breadths lower that the predefined cutoff threshold. Both samples detected above the threshold were positive by laboratory testing, and one of which was an additional detection over no enrichment. Although the samples were not tested for susceptibility to tetracycline, *tetM* was detected in 3 samples before enrichment and five samples after enrichment (4 above and one below the cutoff threshold).

Despite 10-fold representation in final probe library, the AMR genes assessed in this study did not demonstrate a higher coverage depth than the rest of the genome (Figure S2). We also observed uneven enrichment across the target AMR genes, with poor improvements in coverage of the 23S gene after enrichment (median (IQR) coverage depth before enrichment of 1.6 (1.0-3.4) and after enrichment of 6.7 (3.6-12.5)).

### 8.4 Evaluation of multiplexing prior to enrichment

Four samples (80U, 303D, 347UB, 367UB) were multiplexed before hybridisation with the SureSelect probes. The multiplexed run generated 8.2 gigabases (Gb) of sequence data. Once bases from misclassified barcodes and unclassified bases were removed, 1.7Gb were classified as bacterial and, despite enrichment, 1.4Gb classified as human. The median number of bacterial bases classified as *N. gonorrhoeae* was 111 megabases (Mb) (IQR 55-305), with a median *N. gonorrhoeae* single-fold genome coverage of 77% (IQR 67-85) (Table 3). In comparison to no enrichment, multiplexed enrichment achieved a median 12-fold (IQR 4.8-33) improvement in genome coverage depth, which is less than the median 365-fold (IQR 112-720) improvement seen after singleplex enrichment (Table 2). Despite hybridisation of equimolar amounts of barcoded DNA with the SureSelect probes there was uneven coverage across the 4 samples, with >700Mb separating the highest and lowest number of bases classified as *N. gonorrhoeae*.

**Table 3.** Comparison of no enrichment, singleplex enrichment and multiplex enrichment. Total bases classified as *N. gonorrhoeae* (Ngon), single-fold genome coverage breadth, and genome coverage depth reported for the 4 samples sequenced as a multiplex.

|  | Without enrichment |  |  | With singleplex enrichment |  |  | With multiplex enrichment |  |  |  |
| --- | --- | --- | --- | --- | --- | --- | --- | --- | --- | --- |
| Sample | Total Ngon bases | Genome coverage breadth (%) | Average coverage depth | Total Ngon bases | Genome coverage breadth (%) | Average coverage depth | Total Ngon bases | Genome coverage breadth (%) | Average coverage depth | Coverage depth fold-enrichment, multiplex vs no enrichment |
| 80U | 749179 | 3.46 | 7.98 | 1280208433 | 97.44 | 314.12 | 96384278 | 71.91 | 40.05 | 5.02 |
| 303D | 9692745 | 82.96 | 4.73 | 17664552591 | 97.08 | 6637.14 | 1144125623 | 94.61 | 352.47 | 74.5 |
| 347UB | 1066606 | 7.23 | 5.69 | No singleplex data available |  |  | 47875172 | 52.70 | 48688685 | 4.07 |
| 367UB | 1474533 | 12.82 | 4.52 | 6713863610 | 96.63 | 2405.44 | 238397019 | 82.44 | 85.96 | 19.0 |

All 4 samples achieved genome coverage depth of greater than 20-fold (Table 3), which is the minimum depth required for robust detection of AMR determinants(8). Three of the four were resistant to a quinolone (nalidixic acid; 303D, 347UB, 367UB) and one was sensitive (ciprofloxacin, 80U). Depth was insufficient to resolve the sequence of the *gyrA* or *parC* genes in any sample without enrichment, and after multiplexed enrichment adequate depth was achieved to detect mutations in *gyrA* (S91F) in 2/3 resistant samples (303D and 367UB). Sequencing depth was inadequate to determine resistance in sample 347U, with only 7-fold coverage depth of *gyrA*. *blaTEM-1* was detected in samples 303D and 367UB without enrichment, corresponding with the presence of beta-lactamase detected by the laboratory, but was not detected in sample 347UB despite detection by the lab. After multiplexed enrichment, *blaTEM-1* was detected below the cutoff threshold in 1/3 samples reported positive by the laboratory, and in an additional sample without laboratory results (80U).

### 8.5 Assesssment of relatedness and comparison of Nanopore with Illumina sequence data

Comparison of Nanopore (from urine and swab sequencing) and Illumina genomes (from isolate sequencing) in a phylogenetic tree showed concordance between the sequencing methods, demonstrating the ability to infer relatedness: 13/15 (87%) samples had singleplexed enrichment Nanopore data and 11/15 had Illumina data for analysis, giving a total of 11 samples with paired Nanopore and Illumina data (Figure 5). The median (IQR) [range] number of SNPs detected between Nanopore and Illumina sequence pairs generated from matched urine/swab samples and cultured isolates from the same patient was 44 (38–52) [24-245]. This likely represents a combination of sequencing artefact and true differences between the samples.

**Figure 5.**
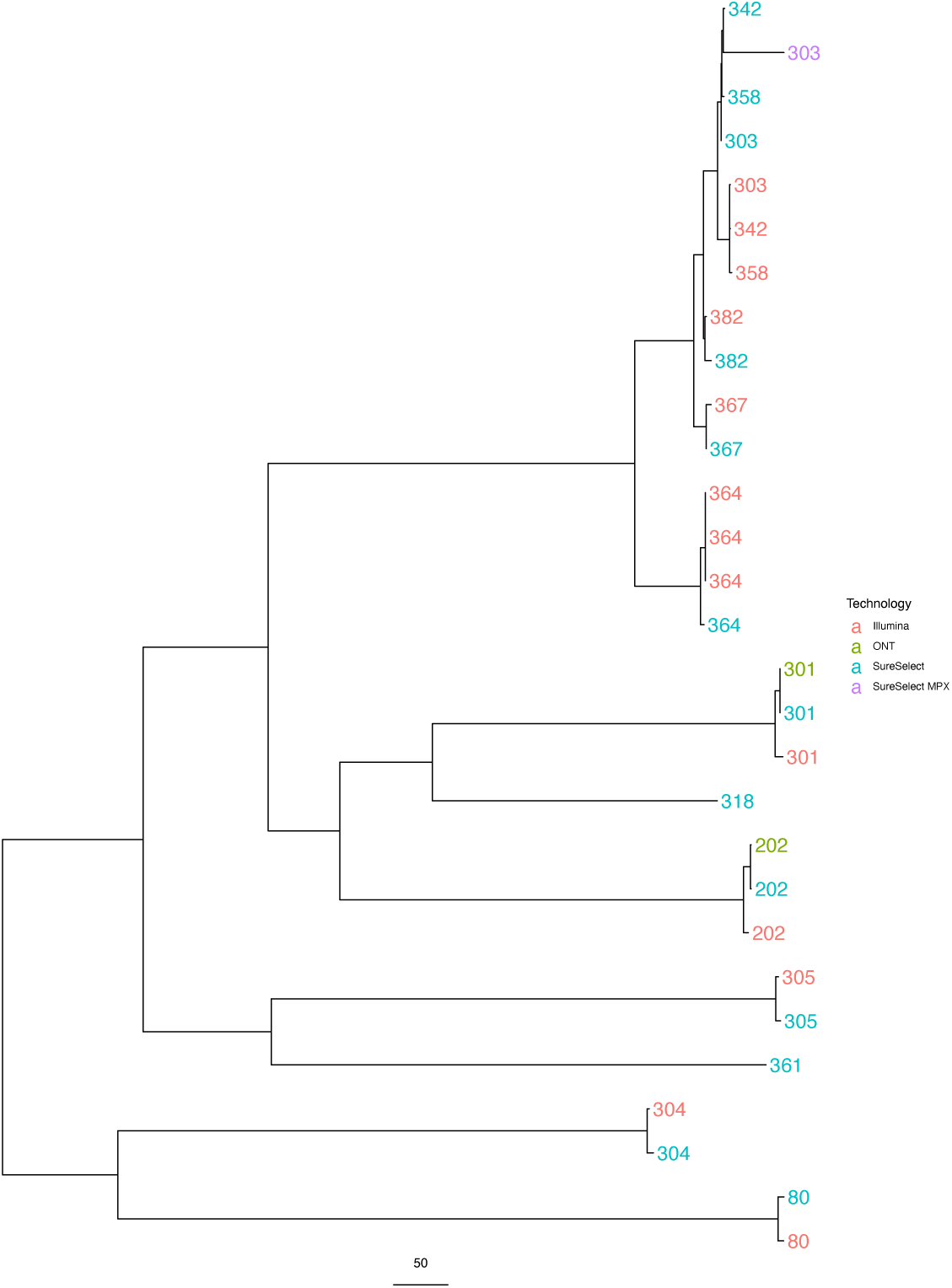
Maximum likelihood tree of genomes generated from consensuses sequences, comparing the same samples sequenced with Illumina (red), Nanopore with no enrichment (green), Nanopore with singleplex enrichment (blue) and Nanopore with multiplexed enrichment (purple). Sample 364 has three Illumina sequences generated from two rectal swab cultures (not included in enrichment and singleplex sequencing) and one urethral swab culture.

Samples genetically further away from the reference genome used showed greater differences between Illumina and Nanopore results, with samples 303D, 342UB and 358UB forming sequencing technology-specific clades instead of grouping by patient, as seen in all the other samples.

Two samples without enrichment (202D, 301D) generated enough genome coverage for inclusion in this analysis and were almost identical to their corresponding SureSelect-enriched pair (2 and 0 SNPs different, respectively). Only one multiplexed sample, 303D, generated enough data to reconstruct the genome for this analysis.

## 9. Discussion

This study demonstrates the utility of target enrichment for the culture-free detection of *Neisseria gonorrhoeae* directly from urine and urethral swabs by sequencing. We also show, for the first time with Nanopore sequencing, that target enrichment can be achieved after multiplexing of samples prior to enrichment.

Where enrichment was successful it was possible to achieve >96.5% genome coverage breadth at a depth of 300-fold or greater, representing a minimum 40-fold improvement in coverage depth. Multiplexing with enrichment still provided >20-fold coverage depth in all samples tested. The depth and breadth of *N. gonorrhoeae* genome coverage achieved after enrichment enabled detection of *gyrA* mutations known to confer resistance to quinolones in all samples with enriched sequence data reported as resistant to nalidixic acid, and additionally detected mutations in a sample with no susceptibility data due to failed growth on culture. Targeted enrichment allowed sequencing at high depth of other chromosomal genes known to confer antimicrobial resistance, e.g., the *mtrR* gene where a mutation at amino acid position 45 confers both multi-drug and macrolide resistance. This offers confidence that resistance should be detected if it were present.

mNGS holds promise for use in routine clinical diagnostics(17) but is hampered by high levels of host DNA, which dominates when nucleic acid is extracted directly from a sample without initial culture, and subsequently overwhelms the sequence data such that information on pathogens is often very limited (for example(18)). Target enrichment enables selection of the genome of interest over contaminating host DNA, and is useful where the target pathogen is known in advance of the diagnostic test. Prevalence of multi-drug resistant *N. gonorrhoeae* and the need for a fast diagnostic that can detect both the pathogen and any corresponding antimicrobial resistance makes targeted sequencing an appealing prospect.

Susceptibility testing relies on culture, but *N. gonorrhoeae* can sometimes be difficult to grow in the laboratory. This study demonstrates the additional value of sequencing with enrichment, detecting *gyrA* mutations with a mean coverage depth of 116-fold in a sample that did not grow in culture.

Detection of plasmid-mediated resistance genes was less successful, with enrichment missing detection of beta-lactamase known to be present by laboratory testing in the majority of samples, and despite detection of *blaTEM-1* in the sequence data of six samples before enrichment. Inefficient amplification of plasmid DNA pre-hybridisation, or the short SureSelect probes not effectively pulling out the plasmid-bound gene during hybridisation, could be possible explanations for the unsuccessful detection of *blaTEM-1* but further work is required to determine the exact reason.

Two samples failed enrichment. Analysis of their post-enrichment sequence data identified predominantly laboratory kit-based contaminants in addition to low numbers of reads mapping to *N. gonorrhoeae*. We propose that this is due to the very low amount of *N. gonorrhoeae* DNA in the original urine extract for these two samples, as has been described by others studying low bacterial load samples(19,20).

Multiplexing allowed a median 12-fold improvement in genome coverage when four samples were barcoded and pooled prior to hybridisation and detection of relevant AMR determinants. Demultiplexing samples after sequencing leads to loss of data as some reads remain unclassified due to a barcoding score lower than the default requirement in Guppy. The larger than expected number of unclassified reads generated from the multiplexed samples could be a consequence of the combination of barcoding and ligation library preparation used to prepare these samples, but further work would be required to determine this. The number of unclassified reads could potentially be reduced by changing the minimum barcoding score requirements, although this carries the risk of increased false-positive calls.

Other limitations of this study include the large starting amount of DNA required for this hybridisation method. Often, DNA extracts directly from clinical samples fail to generate the required 3.5μg required for the ONT Sequence Capture protocol. Extracts from urine and urethral swabs do, in general, achieve the required amount of DNA. However, samples from sterile sites without much host DNA contamination, for example CSF samples, or from samples with a limited volume or size, would not be good candidates for probe-based enrichment with Nanopore sequencing. The method is performed over two days, with a 16-hour overnight hybridisation, which makes it a lengthy assay. Although there could potentially be some optimisation of the protocol, it currently cannot compete with the time it takes to perform a NAAT or other molecular assay, but is still likely faster than culture. There are also limitations arising from artefacts introduced by ONT sequencing, as seen for some samples in Figure 5. However, this study was performed with version 9.4.1 flowcells, and the more recent generation of flowcells, versions 10 onwards, may address this(21). In our data we observed uneven enrichment across target AMR genes, with the 23S gene in particular under-represented in enriched sequence data. Future work could optimise probes in this region of the genome to increase coverage of this important gene.

The nucleic acid amplification tests are a reliable method for detecting *N. gonorrhoeae* but cannot give the whole picture of AMR, an important consideration given the increasing prevalence of multi-drug resistant gonorrhoea. This study demonstrates sequencing with target enrichment can enable high-depth chromosomal antimicrobial resistance gene characterisation and relatedness detection that was not previously possible for these sample types with direct metagenomic sequencing. Additionally, multiplexing prior to enrichment allows higher sample throughput and reduction in the costs associated with both probe-based enrichment and sequencing.

## Data Availability

The sequence data generated in this study are deposited in the European Nucleotide Archive and are publicly available under study code PRJEB64347.

https://www.ebi.ac.uk/ena/browser/view/PRJEB64347

## 12. Author statements

### 12.1 Author contributions

TLS, NDS, ML and DWE designed the study. NDS conducted the bioinformatics analysis. TLS, LB and JK performed laboratory experiments. KC organised clinical samples, performed culturing and collated metadata for samples collected in Brighton. TLS, NDS and DWE prepared the manuscript. The GonFast Investigators Group recruited patients and collected samples. All authors read and contributed to the manuscript.

### 12.4 Ethical approval

This study was conducted with NHS Research Ethics committee approval (reference 19/EM/0029).

## 12.5 Acknowledgements

We thank the microbiology laboratory staff of Oxford University Hospitals NHS Foundation Trust and the University Hospitals Sussex NHS Foundation Trust for providing assistance with sample collection.

The GonFast Investigators Group includes Joanna Rees and Emily Lord (Oxfordshire Sexual Health Service, Oxford University Hospitals NHS Foundation Trust, Oxford); and Suneeta Soni, Celia Richardson, Joanne Jessop, and Tanya Adams (Brighton and Hove sexual health and contraception service, Royal Sussex County Hospital, Brighton).

