## Supplemental Tables S1, S2; Supplemental Figures S1, S2. for "Target enrichment improves culture-independent detection of *Neisseria gonorrhoeae* direct from sample with Nanopore sequencing"

### Supplementary Materials

Table S1. *Neisseria gonorrhoeae* genomes used for SureSelect custom probe library design.

| ***N. gonorrhoeae*** **strain** | **BioSample number** | **BioProject number** |
|---|---|---|
| H18-209/ SRX5491102 (1) | SAMN11081007 | PRJNA523794 |
| NCCP11945 | SAMN02603475 | PRJNA29335 |
| WHO F | SAMEA2448460 | PRJEB14020 |
| WHO G | SAMEA2448461 | PRJEB14020 |
| WHO K | SAMEA2448462 | PRJEB14020 |
| WHO L | SAMEA2448463 | PRJEB14020 |
| WHO M | SAMEA2448464 | PRJEB14020 |
| WHO N | SAMEA2448465 | PRJEB14020 |
| WHO O | SAMEA2448466 | PRJEB14020 |
| WHO P | SAMEA2448467 | PRJEB14020 |
| WHO Q | SAMEA4640835 | PRJEB26560 |
| WHO U | SAMEA2796327 | PRJEB14020 |
| WHO V | SAMEA2796328 | PRJEB14020 |
| WHO W | SAMEA2448470 | PRJEB14020 |
| WHO X | SAMEA2448468 | PRJEB14020 |
| WHO Y | SAMEA2448469 | PRJEB14020 |
| WHO Z | SAMEA2796326 | PRJEB14020 |

Table S2. Genes involved in conferring antimicrobial resistance, represented 10-fold in the final SureSelect custom probe library.

| **Gene** | **Antimicrobial agents** |
|---|---|
| 23S rRNA | Macrolides |
| *gyrA* | Fluroquinolones |
| *ermB* | Macrolides |
| *ermC* | Macrolides |
| *ermF* | Macrolides |
| *mef* | Macrolides |
| *mtrR* | Macrolides, cephalosporins, penicillin, tetracycline |
| *parC* | Fluroquinolones |
| *penA* | Cephalosporins, penicillin |
| *ponA* | Cephalosporins, penicillin |
| *porB* | Cephalosporins, penicillin, tetracycline |
| *blaTEM-1* | Penicillin |
| *blaTEM-135* | Penicillin |
| *tetM* | Tetracycline |

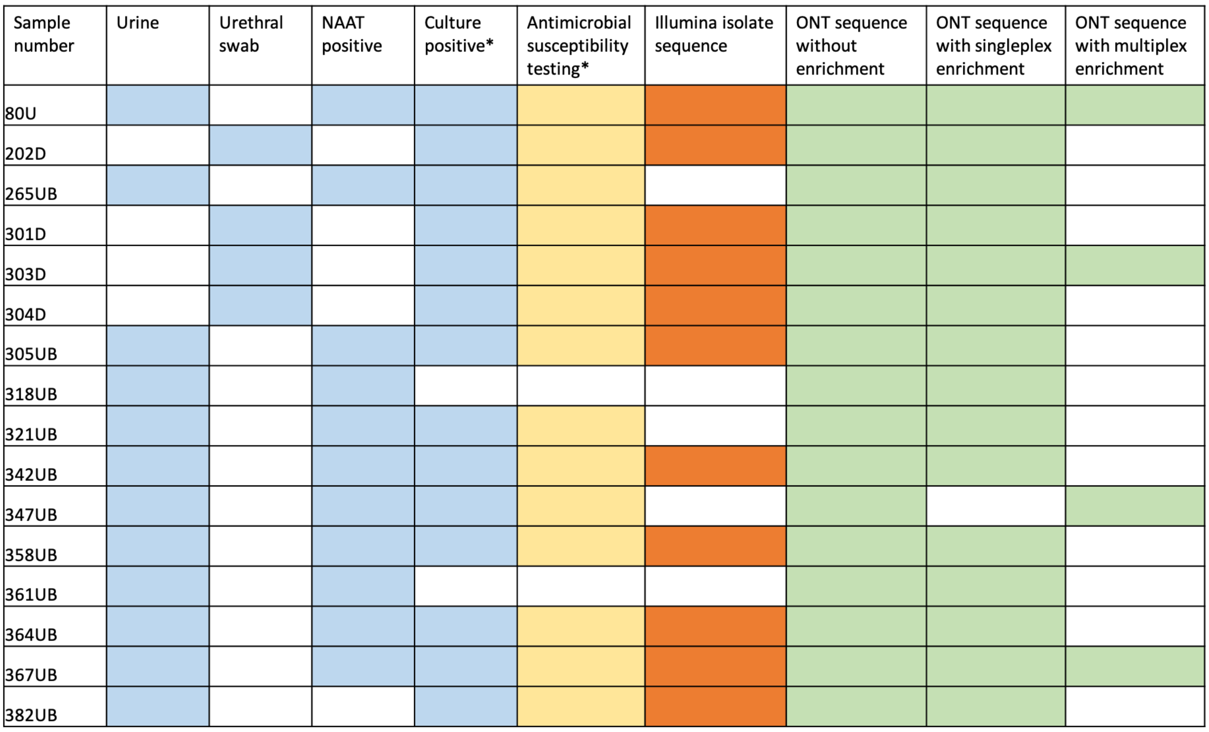


Figure S1. Sample information (blue) and summary of data available for each individual sample: Antimicrobial susceptibility data (yellow); Illumina sequence data from isolate culture (orange); ONT sequence data from urine or urethral swab extract (green). *culture-positive status and antimicrobial susceptibility results reported for urine samples were derived from culture of a corresponding swab collected at the same time as the urine sample.

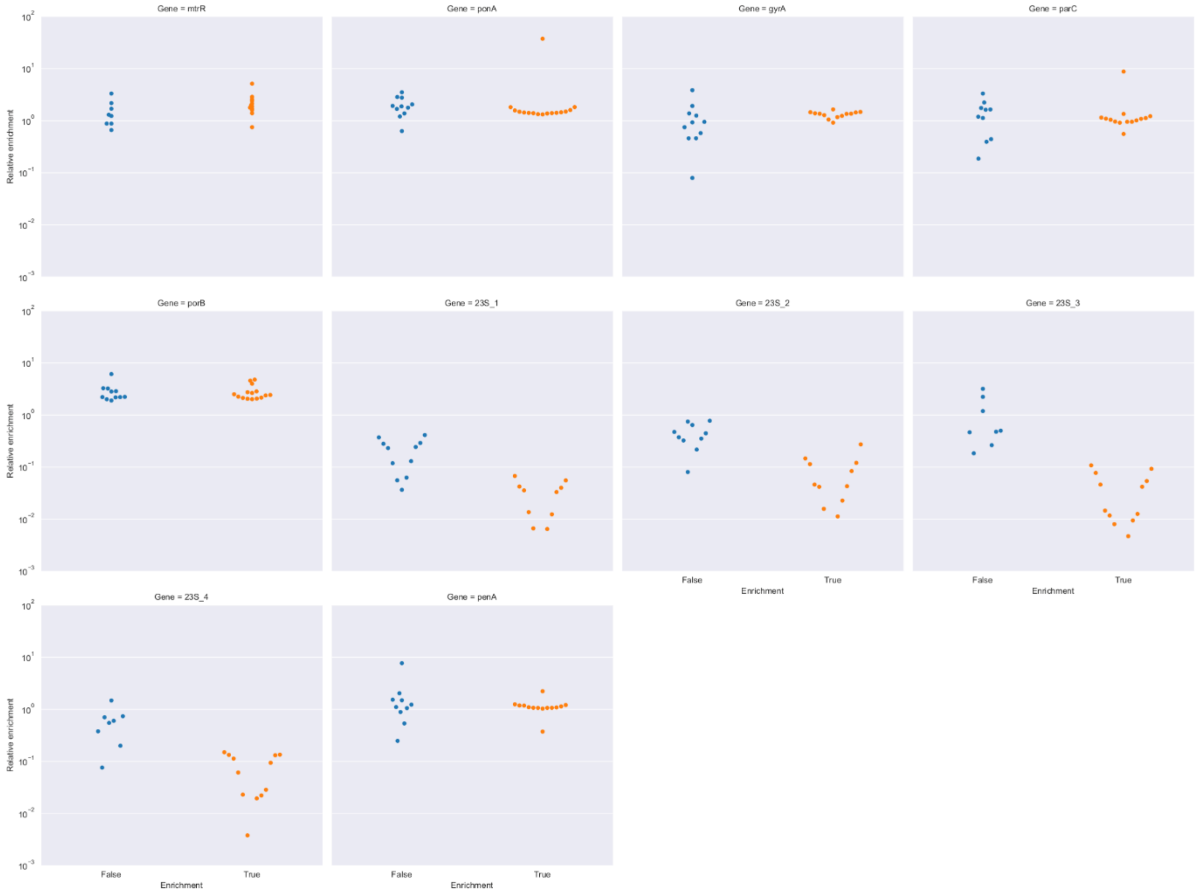


Figure S2. Relative enrichment of AMR genes in comparison to whole genome. Blue represents mean depth of AMR genes relative to mean genome depth before enrichment; orange represents mean depth relative to mean genome depth after singleplex enrichment for 23S, gyrA, mtrR, parC, penA, ponA, porB
